# Multimodal investigation of endometriosis by using LA-ICP-MS and QCL-based infrared microscopy

**DOI:** 10.64898/2026.10.22.26366152

**Authors:** David Hingst, Jessica Weiß, Renata Voltolini Velho, Denny Pellowski, Christoph Schüßler, Stefanie Weigel, Hajo Haase, Sylvia Mechsner, Uwe Karst, Maria Maares

## Abstract

Endometriosis affects millions of women worldwide. It is linked to severe pain and leads to high socio-economic costs. Endometriosis is characterized by the growth of tissue resembling the endometrium outside the uterus, with the cause still unclear. This study demonstrates how multimodal bioimaging methods can provide new insights into the elemental and molecular processes of endometriosis at the macroscopic level.

By using multimodal bioimaging techniques, the role of trace elements (TE) and changes in molecular composition were investigated in human endometriosis biopsies and compared with TE concentrations in serum and peritoneal fluid during the disease. As an element-specific bioimaging technique, laser ablation-inductively coupled plasma mass spectrometry (LA-ICP-MS) was utilized to analyze the TE profile in human biopsies with high sensitivity. Additionally, a quantum cascade laser-based infrared microspectroscopy generated complementary spatial molecular information of the affected tissue. The chemometric processing of the molecular data was made by using uniform manifold approximation and projection (UMAP). To validate the results obtained, TE levels in body fluids were quantified by using ICP-MS and microassays. Our study demonstrates that differences in the TE profile and at the protein level, in terms of mucin expression, can be detected in lesion areas. The TE changes in the tissue differed from those in the body fluids. The presence of mucins in the lesion areas was additionally confirmed by immunohistochemistry. These results highlight the potential of imaging methods in the field of endometriosis research and offer a promising avenue for further studies.

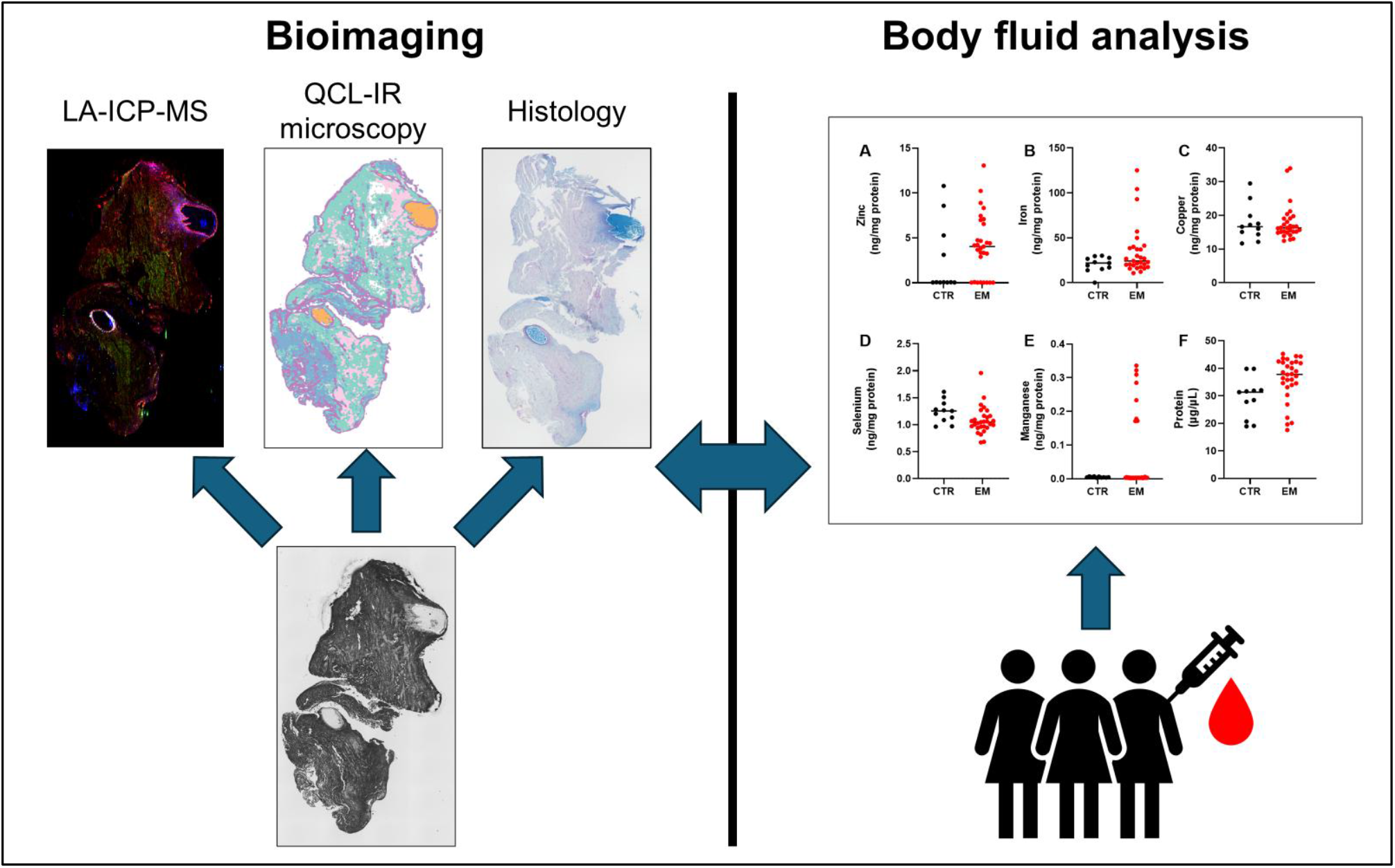

## Introduction

Endometriosis is an estrogen-dependent chronic, inflammatory disorder characterized by endometrial epithelial cells growing outside the uterus.^1,2^ It is usually found in the peritoneum around the pelvic organs, but it can also be infiltrate adjacent organs like intestines or bladder. Symptoms are severe menstrual cramps, strong pelvic pain, especially during menstruation, and heavy menstrual bleeding.^2,3^ There is also a link to infertility in affected women.^4,5^ It is estimated that up to 180 million women worldwide in the reproductive age are affected by endometriosis, which leads to a major social and economic burden.^6,7^ This disease affects a person’s quality of life and causes both physical and emotional discomfort.^8^ Diagnosis remains challenging because the disease is heterogeneous. It can manifest across a wide spectrum of phenotypes and is frequently associated with non-specific symptoms. Historically, the gold standard for diagnosis was direct surgical visualization of lesions, typically via laparoscopy, often with histopathological confirmation. However, current clinical practice increasingly recognizes a non-invasive diagnostic approach that combines clinical assessment with imaging modalities, particularly transvaginal ultrasound and magnetic resonance imaging.^9,10^ To date, no curative treatment is available. Therapeutic strategies, therefore, focus on symptom control and disease management. Hormonal therapy is considered the first-line treatment. It aims to suppress disease activity and alleviate symptoms. Surgical intervention may be indicated, particularly in cases of refractory symptoms or extensive disease, to achieve complete excision of lesions. Additionally, analgesic therapy plays an important role in pain management.^11^

A few studies have already linked endometriosis with dyshomeostasis of essential trace elements (TEs). Particularly, zinc (Zn) seemed to be affected in endometriosis yet with conflicting results, as both decreased levels in blood^12^ and serum^13,14^ were reported, while other studies observed no changes between patients and control group (CTR).^15^ Moreover, supplementing selenium (Se) was found to be promising as an adjuvant therapy for severe pain, a common symptom during endometriosis^16^, while a positive correlation between the occurrence of endometriosis and high Zn intake has recently been discussed ^17^, underlying the possible relevance of these micronutrients for endometriosis pathogenesis. As TE are essential for various physiological functions, including the immune system and antioxidative defense, both of which seem to be impaired during endometriosis^18^, a detailed investigation of (sub)tissue TE-levels during endometriosis and their role in endometriosis progression is needed.

Given the need for rapid, early, and noninvasive diagnosis, as well as a deeper understanding of disease progression, biomedical analytical techniques are opening up new possibilities. Analysis of body fluids such as blood, serum, or menstrual blood could provide a rapid, noninvasive tool for diagnosing endometriosis and can also reveal various molecule classes or element profiles. To investigate the elemental profiles, usually inductively coupled plasma-mass spectrometry (ICP-MS) is used. ICP-MS is a powerful, highly sensitive instrument for analyzing liquid samples and is also used in medical applications. ^12,15^ It is also used to analyze the inorganic composition of affected tissue upon homogenization and acid digestion.

While commonly used techniques, such as ICP-MS, provide total element concentrations, it is difficult to establish correlations between the liquid sample and the elemental distribution within the affected organ or tissue. Biomedical imaging techniques are therefore becoming increasingly important.^19–21^ These techniques can generate spatially resolved information, enabling correlation of the affected tissue area with changes in the elemental or molecular profile. For elemental spatially resolved screening, a laser ablation system is coupled to an ICP-MS, so-called laser ablation ICP-MS (LA-ICP-MS). This hyphenation allows the generation of an image of the elemental distribution of the chosen elements within the tissue section. The high sensitivity of ICP-MS enables the detection of elements at low concentrations like TE, and the external calibration by using matrix-matched standards allows for quantification. So far, ICP-MS has become a widely used method in clinical diagnostics and biomonitoring.^22,23^ Compared with other element-specific techniques, such as micro-X-ray fluorescence, both the spatial resolution and sensitivity are significantly higher when using LA-ICP-MS^24^; however, it is a destructive technique. The LA-ICP-MS analysis does not provide molecular information due to the complete combustion of organic material, resulting in the loss of molecular information. Combining elemental analysis with molecular analysis, such as infrared (IR) (micro) spectroscopy, can therefore be a powerful approach for non-destructive imaging. This multimodal approach improves understanding of the elemental data and provides additional molecular information. Previous research used desorption electrospray ionization mass spectrometry imaging and matrix-assisted laser desorption/ionization mass spectrometry imaging to analyze endometriosis tissue ^25,26^. These approaches, however, are time-consuming and are limited to individual molecular classes. Regarding IR imaging, Fourier transform infrared (FTIR) spectroscopes were often used, but these are limited in scan speed. A further development in the field of IR imaging was the quantum cascade laser (QCL)-based IR spectroscope. These offer spectral coverage over the fingerprint range, which is particularly suitable for biological samples.^27,28^ The fingerprint range contains a lot of biomedical information about proteins, lipids, and carbohydrates. A major advantage of QCL-based IR microscopy is its much faster data acquisition while maintaining spectral and spatial performance comparable to that of conventional FTIR imaging systems.^29,30^

Infrared imaging produces a large amount of data. To interpret the data, versatile chemometric data processing is becoming increasingly important. For the data processing, uniform manifold approximation and projection (UMAP) was used to interpret this high-dimensional data. UMAP is a non-linear dimensionality reduction method that projects high-dimensional data into low-dimensional clusters while preserving both the local and global structures of the original data. Often, the low-dimensional cluster is displayed in 2D. In a 2D representation, the low-dimensional clusters allow individual grouping based on similarity. In the case of infrared imaging data, this means spectral similarity.^31^ The potential of UMAP for elemental imaging and IR screening has already been demonstrated.^21,32^

Thus, this study aimed to elucidate and quantify the spatial distribution of essential TE in endometriosis lesions and to provide insight into the molecular changes in the eutopic and ectopic environment. This work shows for the first time the application of LA-ICP-MS for spatially resolved analysis, combined with spatial molecular analysis using IR-microscopy in affected tissue in endometriosis research, validated with ICP-MS of body fluids, histological staining, and microassays.

### Experimental

#### Samples

The study was approved by the Institutional Review Board of the Charité - University Medical Centre (Ethics approval number EA4/127/23). All patients provided written informed consent. Tissue samples were collected during laparoscopic surgery at Charité – Universitätsmedizin Berlin. Patients were selected based on clinical, intraoperative, and subsequent histopathologic findings. Control samples were collected from women without endometriosis who had undergone laparoscopy for benign gynecological conditions unrelated to endometriosis, such as ovarian cysts, uterine fibroids, hydrosalpinx, pelvic pain, or the unfulfilled wish to have children.

Serum samples were obtained from patients with endometriosis before anesthesia induction on the day of laparoscopic surgery, as well as from outpatients diagnosed with endometriosis at Charité – Universitätsmedizin Berlin. Control serum samples were sourced from the institutional blood donation bank or collected from healthy volunteers recruited via announcements on the Charité intranet and through personal networks. Peritoneal fluid samples were collected intraoperatively during laparoscopy from patients with endometriosis and from women without endometriosis who were undergoing surgery for benign gynecological conditions like ovarian cysts, uterine fibroids, hydrosalpinx, pelvic pain, or infertility. They were aspirated from the Douglas pouch immediately after trocar insertion to minimize contamination with blood. All samples were immediately processed, aliquoted, and stored at −80 °C until analysis. Table 1 summarizes the number of samples and ages of the patients involved in all the TE quantification analyses.

**Table 1.** Overview of patients, serum, and peritoneal fluid samples in this study.

|  | Serum |  | Peritoneal fluids |  |
| --- | --- | --- | --- | --- |
|  | EM | CTR | EM | CTR |
| **N** | 37 | 44 | 29 | 12 |
| **Age (median, IQR)** | 31.0 (26.0; 34.0) | 25.0 (22.0; 30.0) | 31.0 (26.5; 36.0) | 34.0 (30.5; 34.0) |

IQR, interquartile range. EM, endometriosis patients; CTR, controls

#### Sample preparation

The endometriosis Research Center Charité provided formalin-fixed and paraffin-embedded samples. Five Samples of peritoneal endometriosis (PEM) and five samples of deep infiltrating endometriosis (DIE) were selected. Additionally, five location-matched control samples were included for each subtype: five samples of peritoneum and one sample each of rectum, ovary, and intestine were prepared. The Samples were cut into 5 µm thin sections using HistoCore AUTOCUT – Automated Rotary Microtom (Leica Biosystems, Hußloch, Germany) and mounted on SuperFrost® Plus microscope slides (Epredia, Breda, the Netherlands) for the LA-ICP-MS analysis. To analyze the sample using IR microscopy, the samples were cut into 5 µm sections and mounted on indium tin oxide-coated glass slides (Diamond Coatings, Brierley Hill, UK).

For the initial analysis, the samples were deparaffinized and rehydrated by the following steps: two times xylene (Merck KGaA, Darmstadt, Germany) (3 min each), two times 100 % Ethanol (Avantor, Radnor, PA, USA) (1 min each), 95 % and 70 % Ethanol (1 min each) and two times doubly distilled water (3 min each).

Optical images of the washed samples were made with a BZ-9000 microscope (Keyence, Osaka, Japan).

#### Laser ablation-inductively coupled plasma-mass spectrometry analysis

For LA-ICP-MS analysis, a laser ablation system with a wavelength of 266 nm (imageBIO266, Elemental Scientific Lasers, Bozeman, MT, USA) was coupled to an iCAP TQ ICP-MS (Thermo Fisher Scientific, Bremen, Germany) using a dual concentric injector (DCI, Elemental Scientific Lasers, Bozeman, MT, USA).

To identify the elemental profile of the deparaffinized tissue, endogenous elements were detected and recorded as [^31^P^16^O]^+^, [^56^Fe^16^O]^+^, ^65^Cu^+^, and ^66^Zn^+^ using oxygen as the reaction gas with a dwell time of 25 ms per element. The ICP-MS was tuned daily. Ni sampler cone and skimmer cone were chosen for the interface. The plasma power was set to 1550 W, with auxiliary and cool gas flows of 0.80 and 14 L/min, respectively. The laser ablation was performed by using a spot size of 15 μm with a scan speed of 150 μm/s and 1 J/cm^2^ laser energy with a frequency of 100 hz. The ablated sample was transferred with a helium gas flow of 1000 mL/min into the ICP-MS.

Ten matrix-matched standards made of gelatine were prepared to quantify the concentration of Fe, Cu, and Zn by using an external calibration. Each of the elements was spiked to the gelatine standard in different concentrations in a range of 1 µg/g to 1000 µg/g. The elements were added as the following salts: iron sulfate, copper sulfate, and zinc chloride. Standards were cut in the same thickness of 10 μm as the samples using Cryostar NX70 cryostat (Thermo Fisher Scientific, Bremen, Germany). The limit of detection (LOD) was 1.6 µg/g, and the limit of quantification (LOQ) was 5.9 µg/g for iron. LOD of 3.0 µg/g and LOQ of 9.9 µg/g for copper and LOD of 1.4 µg/g and LOQ of 5.9 µg/g for zinc was reached. The LODs and LOQs were calculated using the 3σ and 10σ criteria.^33^

The in-house-developed software Imajar 3.64 (written by Robin Schmid) was used to process the images for the localization and quantification of the elements in the section.

#### QCL-based infrared microscopy

QCL-based infrared imaging was carried out with the Hyperion II-ILIM (Bruker Optics, Ettlingen, Germany) on parallel sections. Imaging scans were operated by using the OPUS software (9.0.0.303) in (transflection) reflection mode with a 3.5x IR objective. Spectral parameters were the following: spectral resolution of 8 cm^-1^, step size of 4 cm^-1^ in a wavenumber range of 950 cm^-1^ to 1800 cm^-1^.

The image processing and UMAP clustering were utilized by the self-developed software Multiscale-Image-Analysis (version 1.0.6) of Hennes Rave.^34^ OriginPro® 2024 (OriginLab Corporation, Northampton, MA, USA) was used to visualize the spectra.

#### UMAP parameters

For UMAP clustering, parameters such as the number of neighbors, minimum distance, and metric can be adjusted, as described by Kronenberg et al.^3^. Each sample was clustered twice. The first cluster was calculated in an euclidean metric to separate the background pixel from the sample pixel. The euclidean metric is more focused on the difference in intensities and is therefore well-suited for distinguishing the sample pixels from background pixels. The second cluster was performed in a cosine metric to highlight the spectral differences. The cosine metric can be used to show more detailed spectral differences.

The number of neighbors was set to 15 to maintain the local structure of the cluster. Minimum distance was set to 0.1 to generate tightly packed clusters and show the topological structure of the clusters. Neither parameter was changed between the first and second clustering. The Leiden algorithm was used to ensure reproducible cluster coloring. It was applied to the second embedding. The resolution parameter was set to 0.18 for all measurements.

#### Quantification of trace elements in body fluids

For the quantification of the TEs Mn, Fe, Cu, Zn, Se, and I in human serum, a validated multi-element method based on ICP-MS/MS was applied.^35^ Thawed samples were diluted 1:10 with a solution containing 5 vol.% 1-butanol (Alfa Aesar, Karlsruhe, Germany), 0.05 m% Na-EDTA (Triplex® III, pro Analysis, Merck, Darmstadt, Germany), 0.05 vol.% Triton™ X-100 (10% in H₂O, Merck-Sigma Aldrich, Steinheim, Germany), and 0.25 vol.% ammonium hydroxide (25% in H₂O, Fluka, Buchs, Switzerland) in deionized water. External calibration standards (for all elements except Se) were prepared from 1000 mg/L single-element stock solutions (Carl Roth, Karlsruhe, Germany). Rhodium (1 µg/L) and Germanium (10 µg/L) were diluted from a 1000 mg/L single-element stock solution (Carl Roth, Karlsruhe, Germany) and added as internal standards. Quantification of Se was performed by isotope dilution analysis using an internal standard solution containing 10 µg/L Se-77 (certified by Trace Sciences International, Ontario, Canada; purchased from Eurisotop SAS, Saarbrücken, Germany). The resulting mixtures were directly analyzed by ICP-MS/MS (Agilent ICP-QQQ-MS 8800, Agilent Technologies, Waldbronn, Germany). In each analytical run, certified reference sera (ClinCheck® Serum Level 1 and 2, RECIPE Chemicals + Instruments, Munich, Germany; and Seronorm™, Sero AS, Billingstad, Norway) were included to ensure analytical accuracy. Blank samples (deionized H₂O) were processed in parallel to determine the LOD and LOQ.

For the quantification of the TEs in peritoneal fluid samples, 500 µL of each sample were digested using microwave-assisted acid digestion (Mars6, CEM, Kamp-Lintfort, Germany). The reference materials used for validating complete microwave digestion were fish muscle (ERM®-BB422, European Commission, Joint Research Centre, Geel, Belgium) and pig kidney (ERM®-BB186, European Commission, Joint Research Centre, Geel, Belgium). Subsequently, a further 1:5 dilution was performed before the TEs in the peritoneal fluids were quantified by ICP-MS/MS as described above.

In peritoneal fluids, 12 CTR samples and 22 endometriosis samples were below the LOQ for Mn, and 7 CTR and 8 endometriosis samples for Zn. To enable a comparison between CTR and endometriosis peritoneal fluid in case of left-censored data, where the measured TE was <LOQ, the lower bound approach was applied^36^. Accordingly, censored values were substituted by LOQ/√2 for values less than LOQ. (2). Finally, the TE concentration was normalized to protein content of the samples, which was analyzed using a homogenized sample aliquot with bicinchoninic acid (BCA)-assay as described^37^. Free Zn as a biomarker for Zn status in serum was quantified following the method from Alker et al.^38^ using the zinc-responsive low molecular weight fluorescent sensor Zinpyr-1 (Santa Cruz Biotechnology, Dallas, TX, USA).

#### Structural analysis for the localisation of lesions and histochemical analysis of Glycoproteins

Tissue sections were first deparaffinised with xylene and then rehydrated with a series of ethanol solutions. Slides for the analysis of glycoproteins were then incubated in alcian blue (AB) (pH 2.5, #101647, Sigma-Aldrich), which stains acidic glycoproteins at low pH, for five minutes at room temperature, after which they were thoroughly rinsed in distilled water. Counterstaining of nuclei was then performed using nuclear fast red (N069.1, Carl Roth). To identify endometriosis lesions, tissue sections were incubated with haematoxylin, which stains cell nuclei and is used in the clinic for tissue structure analysis of endometriosis biopsies, for 10 seconds and incubated for another ten minutes in running tap water, after which slides were thoroughly rinsed in distilled water. Following a process of dehydration through a graded ethanol series, the tissue slices were mounted using a permanent mounting medium. The slices were examined under a light microscope model KEYENCE BZ-X810 (Keyence GERMANY GmbH, Neu-Isenburg, Germany), and images were captured for qualitative and quantitative analysis.

## Results and Discussion

### Elemental bioimaging of endometriosis lesions with LA-ICP-MS

Figure 1 shows the elemental distribution of the elements phosphorus (P), Fe, Cu, and Zn. P is commonly analyzed in LA-ICP-MS studies as it well represents the tissue’s structure and homogeneity.

**Figure 1.**
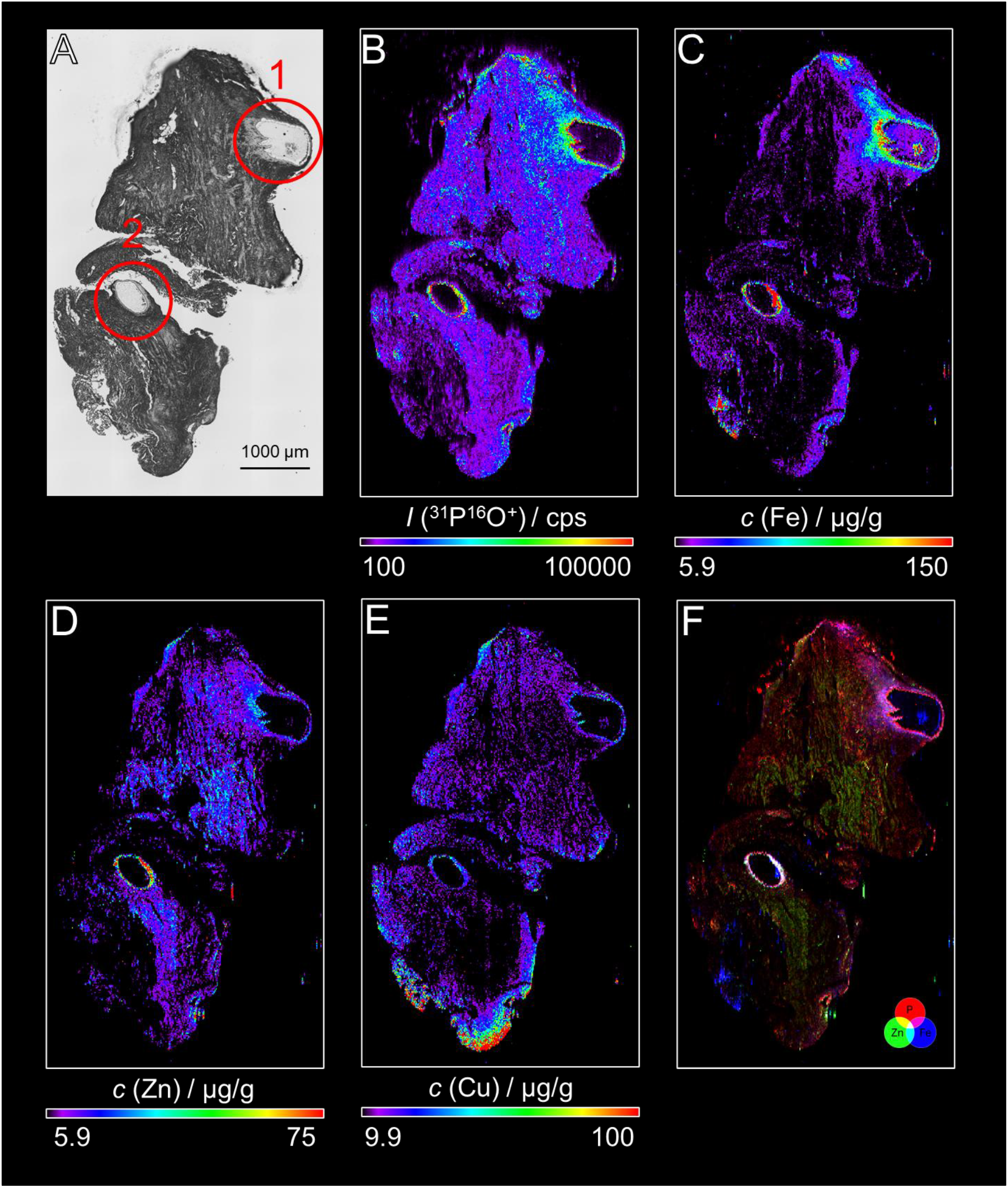
TE distribution in endometriosis lesion. A) Brightfield microscopic image of peritoneal endometriosis lesion (sample EM440), with red marks to highlight the lesions. B) Elemental images obtained with LA-ICP-MS are shown for P (B), Fe (C), Zn (D), endometriosis (EM) lesions are small microenvironments with a typical layer of epithelial, stromal and smooth muscle cells with their own blood vessel and nerves fibers system. Moreover, lesions are often filled with blood and cellular fluids, and macrophages and other immune cells are present due to inflammatory reactions and the elemental image of Cu (E). F) overlay image of P, Fe, and Zn.

Preliminary analyses of endometriosis tissue samples indicated that Mn and Se signals were below the limit required for reliable imaging. Figure 1A showed a brightfield microscope image of the endometriosis biopsy. The red circles mark the lesions in this section. Endometriosis lesions were identified using hematoxylin staining (SI Figure 1). Endometriosis lesions are small microenvironments with a typical layer of epithelial, stromal, and smooth muscle cells, with their own blood vessels and nerve fiber systems. Moreover, lesions are often filled with blood and cellular fluids, and macrophages and other immune cells are present in response to inflammation.^39^ Figure 1B visualizes the distribution of P. In the lesion region, the P signal intensity was much higher than in unaffected areas and correlated well with the thick layer of epithelial and stromal cells. For lesion 1, a gradual increase in signal intensity towards the lesion was also observed. However, this tapering was not evident in lesion 2, although the signal intensity surrounding of the epithelial cell layer is much higher than in lesion 1. Figure 1C shows the distribution of Fe. Similar to the P-distribution, the iron concentration increases around lesions 1 and is particularly high in the epithelial cell layer for lesion 2. It can therefore be assumed that blood and/or Fe-containing cellular matrix are inside lesion 1. This trend is also observed for Zn (see Figure 1D).

While the Zn concentration did not increase much in lesion 1, it was much higher in lesion 2 than in the unaffected areas. Unlike the other elements, Cu showed almost no change around lesions 1 and 2, with only a small increase in the epithelial cell layer. Quantification of TE was performed using gelatin sections containing different concentrations of TE standards (Figure 2). Overall, this results in a mean Fe concentration of 49 µg/g within the lesion 1 region, 203 µg/g within the lesion 2 region, and 15.0 µg/g within the rest of the tissue. The increase in TE concentration is also observed for Zn, with a mean tissue concentration of 7 µg/g, an increase of lesion 1 to 12.8 µg/g and 22.6 µg/g for lesion 2, respectively. For Cu, the increase is less pronounced. On average, the tissue contained 12.5 µg/g Cu, 16.1 µg/g for lesion 1, and 16.5 µg/g for lesion 2, respectively.

**Figure 2.**
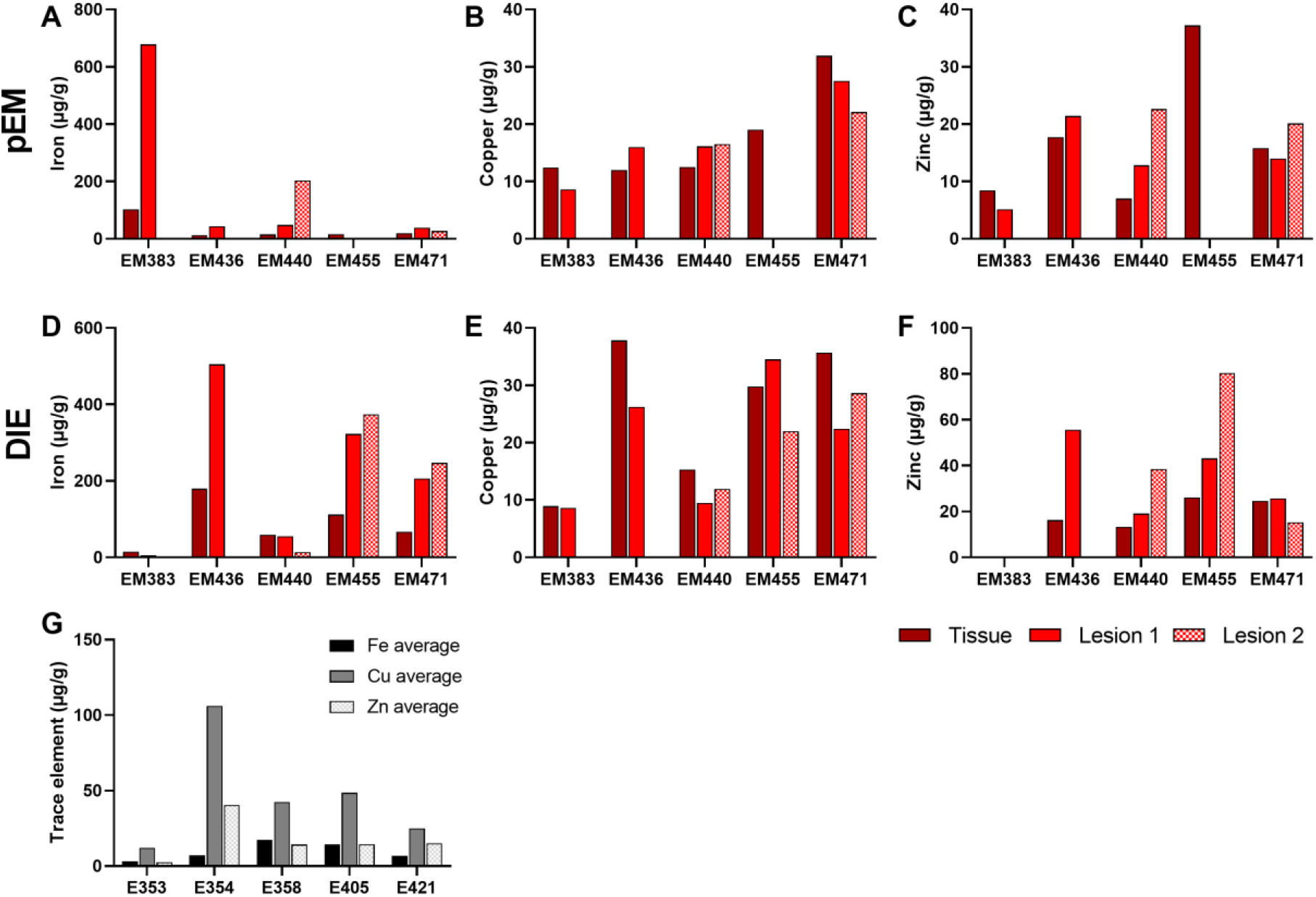
TE concentration in lesions and tissue. Shown are the averaged TE concentrations in peritoneal endometriosis, N=5 (A-C) and DIE (D-F). For this, TE concentrations were measured with LA-ICP-MS, N=5 regions of interest (ROIs) were randomly selected to measure the concentration of Fe (A, D), Cu (B, E), Zn (C, F), and EM and controls (G). Data are presented as mean of N = 5 different EM samples and N=5 control samples.

To highlight the correlation between the increase in TE in the lesion environment, consisting of stromal cells and the ectopic endometrial epithelial cells, an overlay of the distribution of P, Fe, and Zn was shown in Figure 1F. Since Cu showed no significant changes in concentration in the affected areas, it was excluded from the overlay. Each element was assigned a color (P was assigned red, Zn to green, and Fe to blue), and the color intensity indicates high or low concentrations. When several elements were present, the pixel was displayed in a mixed color (as shown in the color overlay diagram), whereas the co-localization of all three elements resulted in a white pixel (Figure 1F). Accordingly, Fe and P were co-localized in lesion 1, and all three elements co-localized in lesion 2. As described in the literature, the endometriosis lesion is characterized by a high expression of endometriotic epithelial cells.^39,40^ The presence of a higher cell density, caused by an increased number of epithelial cells, would fit with the observation of increasing P signal intensities.

The discussed changes in TE profile were also observed in additional samples with a different type of endometriosis (SI Figure 2-4). In total, we analyzed 5 samples from PEM, DIE, and the control group. The measured tissue concentrations of Fe and Zn varied from 38.3 µg/g up to 678.6 µg/g for Fe and from 12.8 µg/g to 80.3 µg/g for Zn within the lesion area. There was also a correlation of Fe and P in the affected tissue and surrounding the lesions. In contrast, the TE profile in the control sample was evenly distributed throughout the tissue with barely any change in Fe concentration and Zn. Changes of TE profiles in the affected tissue could indicate an altered homeostasis during endometriosis. A dyshomeostasis of zinc and iron was evident primarily within the tissue, particularly in the area of the lesion.

It is known that blood and endometriotic cells can accumulate in the lesions during menstruation and remain there, which may explain the increased concentration of iron within the cell-free space.^41^ In addition, high concentrations of Fe and Zn in affected inflammatory tissue could indicate the presence of macrophages, as both play important roles in their homeostasis.^42–44^ It is well known that macrophages are present in endometriosis tissue and play an important role in inflammatory reactions.^45^

### Characterization of the molecular environment of endometriosis lesions with QCL-based IR-imaging

Figure 3A showed the average spectra of each colored segment of the UMAP embedding. The segmentations and coloring were done manually in the software. In Figure 3B the corresponding cluster of the UMAP embedding was shown. After embedding and segmentation, each spectrum and its corresponding pixel were colored according to the segment in which the spectrum clusters. It visualized the UMAP cluster using a false-color map (Figure 3C). UMAP segmentation was also performed for the DIE sample (SI Figure. 3) and the control sample (Figure 3D-F).

**Figure 3.**
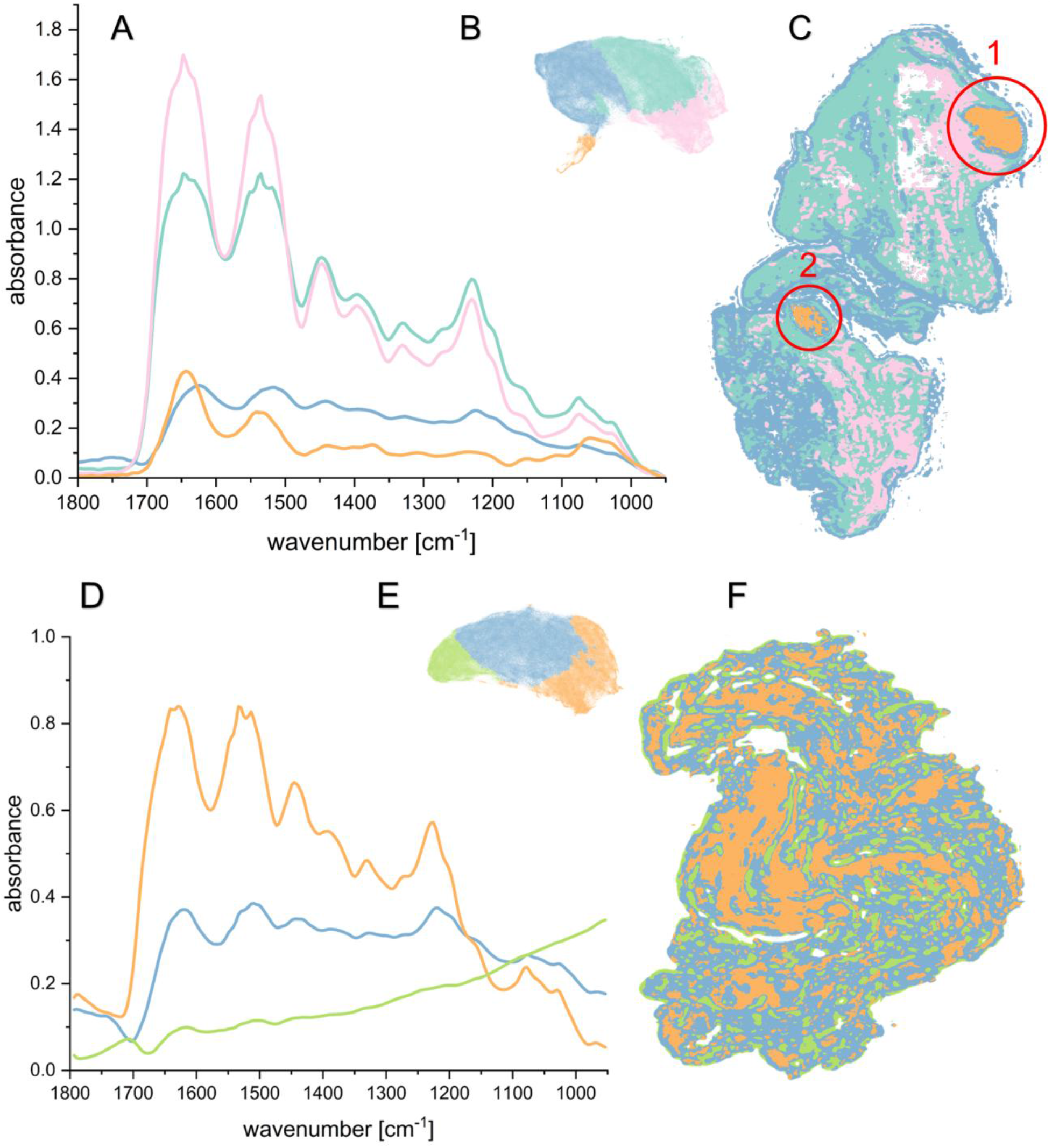
IR-fingerprint of an EM lesion. A) Average spectra of individual-colored segments of the UMAP embedding of the peritoneal endometriosis sample (EM440). B) UMAP embedding of the infrared image data of the peritoneal endometriosis sample. The parameters were set as follows: n_neighbours = 15, min_dist = 0.1 in a cosine metric. The Leiden algorithm was used to color the segments with a resolution parameter of 0.18. C) IR image, colored according to the segments of B. D) Average spectra of individual-colored segments of the UMAP embedding of the control sample (358). E) UMAP embedding of the infrared image data of the control sample. The parameters were set as follows: n_neighbours = 15, min_dist = 0.1 in a cosine metric. The Leiden algorithm was used to color the segments with a resolution parameter of 0.18. F) IR image, colored according to the segments of E.

The averaged spectra of each segment in Figure 3A revealed differences in absorbance of characteristic bands: Accordingly, the pink segment indicated the highest absorbance for the amide I band (C=O stretching, N-H bending, and C-N stretching) at 1650 cm^-1^ and the amide II band (N-H bending and C-N stretching), which both represent protein-specific bands. The band at 1450 cm^-1^ (CH2 and CH3 deformation modes) as well as the amide III band at 1334 cm^-1^ (N-H bending, C-N stretching, and CR-H bending) also pointed to protein. Furthermore, the turquoise segment showed high absorbance across all protein-specific bands. In addition, the characteristic DNA bands at 1230 cm^-1^ (PO^2-^asymmetric stretching) showed a high absorbance, although the absorbance of DNA bands of the turquoise segment is higher compared to the strong protein bands. In the carbohydrates area at 1150 cm^-1^ to 1000 cm^-1^, the absorbance of the glycogen-specific bands 1150 cm^-1^ (C-O stretching), 1078 cm^-1^ (C-C stretching), and 1030 cm^-1^ (COH deformation) were lower for the pink compared to the turquoise and blue segments. This suggested that the pink segment represents a region of tissue with higher protein levels and fewer carbohydrates. The turquoise segment presented tissue with higher carbohydrate levels and slightly lower protein. Both segments were located in regions of lesions 1 and 2. The blue segment is located at the edge of the tissue, where the pixel is filled with tissue and ITO slide, so-called mixed pixels, and also represents tissue with low absorbance (Figure 3B).

The pink, turquoise, and blue segments merge to form a large cluster, while the orange segment is separated from it (Figure 3B), indicating that the segments exhibit clear differences in their spectra. Interestingly, this distinct orange segment represented tissue within the lesions (Figure 3C). The averaged spectra of the orange segment showed lower absorbance across the protein-specific bands but much higher intensities for the carbohydrate area at 1060 cm^-1^ and 1030 cm^-1^. Moreover, the absorbance band for the orange segment was slightly shifted to lower wavenumbers, where absorption at 1060 cm^-1^ (C-O stretching) could indicate the presence of mucins inside the lesion^46^. These glycans are components of the extracellular matrix (ECM), and alterations in ECM composition are known to influence cell properties. This has also been shown in the context of endometriosis.^47^ Additionally, alcian blue staining was performed to verify the presence of carbohydrates, specifically from glycoproteins (SI Figure 6). The staining showed a high accumulation of glycoproteins inside and surrounding the lesions and the orange segment. Similar results can be seen for the staining of the DIE sample (SI Figure 7). This aligns with previous observations that endometrial epithelial cells overexpress mucins, particularly MUC1, which was additionally associated with elevated glycosylation in endometriosis lesions.^48^ In contrast, the control sample (Figure 3-F) shows only a compact cluster and homogeneous segmentation across the entire sample. The absorption of the specific bands is also much lower than in the affected samples.

### Quantification of trace elements in biofluids of endometriosis patients

To gain further insight into the TE status of endometriosis patients and validate the obtained bioimaging data, essential TE were quantified in the serum of endometriosis patients (Figure 4. A-I). Cu, Fe, and I concentrations did not differ between CTR and endometriosis groups and fell within the reference range for these TE (reference ranges Cu 690-2000 µg/L^49^; Fe 614 – 1396 µg/L^50^, I 40 – 80 µg/L^49^; Figure 4A-C). Even though no significant differences were observed in Zn status of CTRs and patients, 12 patients (32%) fell below the cut-off for Zn-deficiency of 642.5 µg/L^51^, whereas only 4 (9%) of CTR could be considered Zn deficient (Figure 4D). In contrast, free Zn is significantly higher in the serum of endometriosis patients (Figure 4E). This Zn species represents the readily available zinc species in the blood and has been discussed as a biomarker for changed Zn homeostasis.^38^

**Figure 4.**
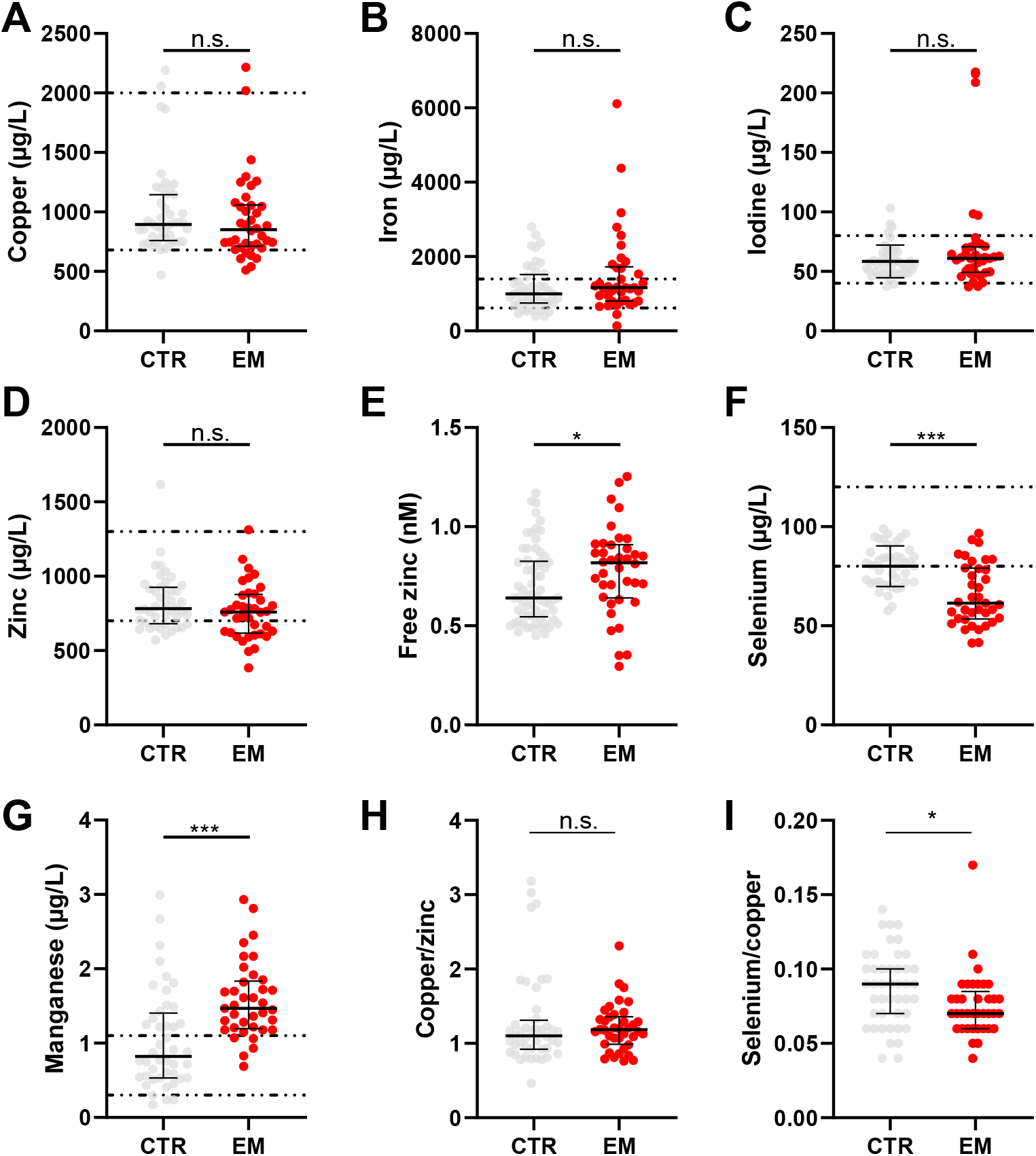
TE concentration in serum samples. Data is shown as median with interquartile range, including the individual values of CTR: N = 12; EM: N = 29). Dashed lines represent the reference values for the respective TE, if available. Significant differences between CTR and EM were tested with Mann-Whitney test (* p <0.05; ** p < 0.01; *** p <0.001).

Se status showed the most pronounced alterations in the cohorts investigated in this study: 66% of the endometriosis patients and 43% of CTRs had Se serum concentrations below the reference range (from 80-120 µg/L^52^) while 14 patients (32%) significantly lower than the CTR group (median EM = 61.0 µg/L vs. CTR = 80 µg/L; Figure 4F). Mn concentrations were mainly above the reference range for serum (0.3 – 1.1 µg/L^49^; Figure 2G), yet comparable to earlier human intervention trials^53^. Most importantly, Mn concentrations were significantly higher in endometriosis patients than in CTRs (median EM = 1.47 µg/L vs. CTR = 1.04 µg/L). Lastly, while the Cu/Zn ratio did not differ, the ratio of Se and Cu was significantly lower in the serum of endometriosis patients (Figure 4I).

In addition to serum of endometriosis patients, we screened TE concentrations in peritoneal fluid, which is produced in the peritoneum, covers all abdominal organs, and ensures communication within the peritoneal microenvironment ^54^. This serous fluid is analyzed in the diagnosis of abdominal diseases, cancer, and gynecological diseases, such as endometriosis, as it contains cytokines, immune cells, and inflammatory markers ^54,55^. Even though there are no significant differences in the amount of TE in CTR and EM PF (Figure 5A-E), Mn concentration in the fluids of 27% of the patients were considerably higher than in CTR PF, where all measured samples were lower than LOQ. Se again tended to be lower in peritoneal fluids (Figure 5D), and Zn showed a trend to increased concentrations in PF (median (CTR) = 0.07 ng/mg protein vs. median (EM) = 4.04 ng/mg protein; Figure 5A).

**Figure 5.**
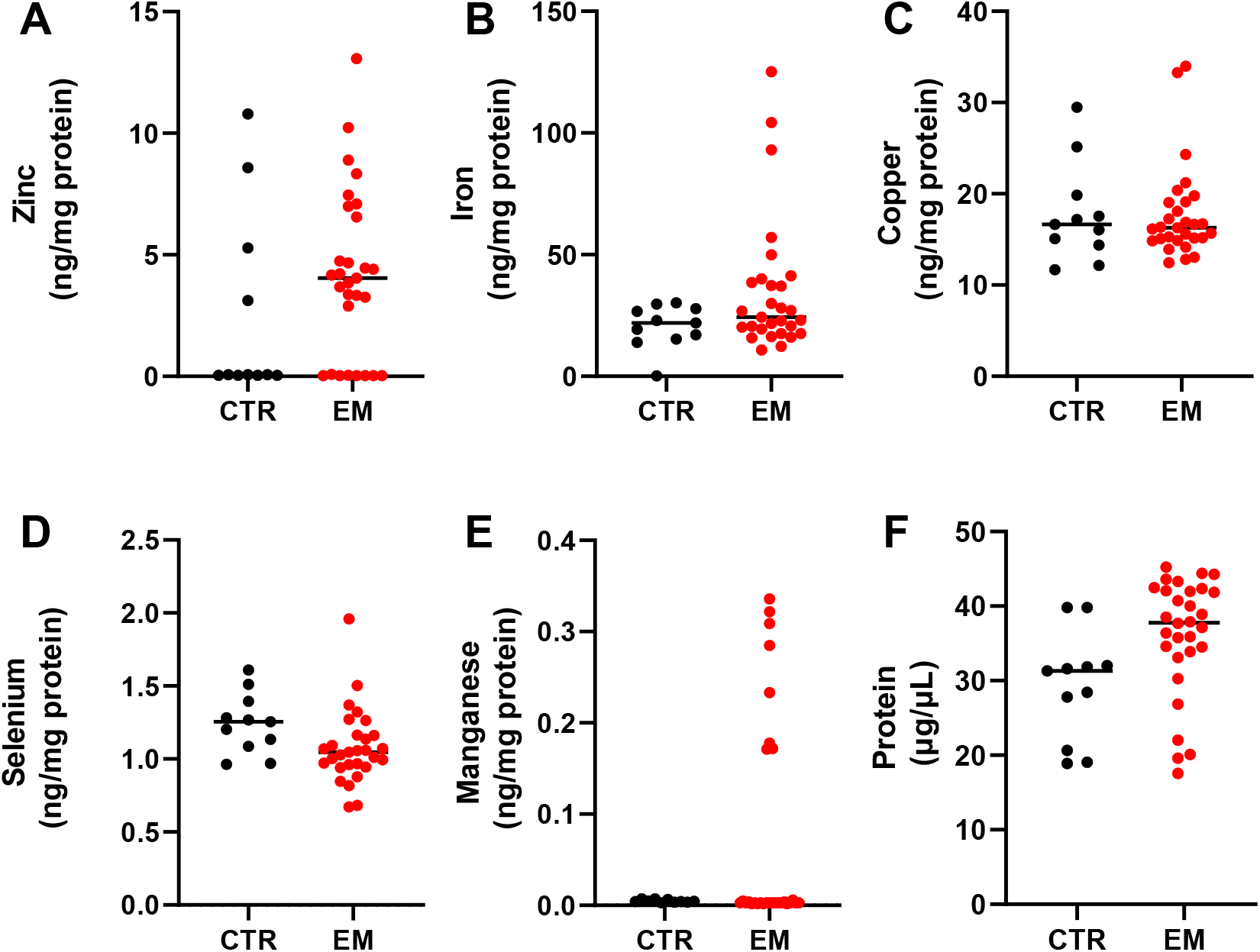
TE concentration in peritoneal fluid. The concentration of zinc (A), Iron (B), copper (C), selenium, and manganese (E) in peritoneal fluids was measured with ICP-MS/MS and depicted relative to the protein content of the sample. The variation of protein concentration in the fluids is presented (F). Shown are the individual values, including the mean and standard deviation of CTR: N = 12; EM: N = 29). Significant differences between CTR and EM were tested with Mann-Whitney U test.

TE screening in serum and peritoneal fluid illustrated pronounced differences in the TE status of CTRs and endometriosis patients, particularly for Se, Mn, and Zn. Low Se is associated with impaired immune defense, redox-imbalance, increased susceptibility to inflammation^56^, and also infertility ^57^, all characteristics of endometriosis ^18^. In addition, elevated Mn concentration was linked to higher oxidative stress and is discussed to be a critical factor for inflammatory conditions or diseases ^58^ As free Zn is pivotal for extra- and intracellular signaling, thereby modulating immune reactions and inflammatory processes^59^ alterations in free Zn could be related to the unbalanced immune system during endometriosis^60^. In summary, TE quantification in biofluids indicated insufficient status or dyshomeostasis of some essential TE, which could be involved in the etiopathogenesis or progression of endometriosis. The altered TE profile of body fluids during endometriosis complements the bioimaging data, indicating a Zn and Fe dyshomeostasis within the tissue and lesion environment, underlying the importance of multi-approached metal analysis in tissue and biofluids.

### Conclusions and Outlook

This work highlights the potential of combining QCL-based IR microspectroscopy with UMAP segmentation for data processing and LA-ICP-MS in the research of endometriosis for spatial analysis. Since the exact cause of endometriosis remains unclear and there are very few treatment options available, it is essential to deepen our understanding of the disease, including its underlying molecular mechanisms. While the analysis of serum and peritoneal fluid from patients with and without endometriosis focused on TE, spatially resolved analysis enabled the detection of elemental and molecular profiles. This approach is highly multimodal and shows potential for obtaining insights into the affected tissue areas.

In endometriosis, TE homeostasis appears to be altered, as evidenced by TE distribution within endometriosis lesions and quantitative TE analysis of serum and peritoneal fluid. LA-ICP-MS showed increased Zn, Fe, and P around the lesions. Interestingly, concentrations measured in serum were lower (Zn, Fe) than those within the lesions. Additionally, while biofluid analysis mostly indicated alterations in Se and Mn homeostasis in endometriosis, TE tissue distribution revealed a spatial increase of Zn and Fe within the endometriosis lesion area. In addition to the LA-ICP-MS-generated elemental information, infrared microscopy was used to map and characterize molecular changes within the endometriosis lesion environment. After data processing with UMAP embedding, it became possible to segment the sample into six clusters. Clusters around and within the lesions showed differences in protein and carbohydrate levels compared with unaffected areas. This work demonstrates that multimodal analytical chemical methods can improve endometriosis research and provide valuable insights into the disease.

## Data Availability

All data produced in the present work are contained in the manuscript.

## Acknowledgement

We thank all patients who took part in this study. Your willingness to donate samples and share your time made this work possible. We are grateful for your contribution.

This work was funded by the Deutsche Forschungsgemeinschaft (DFG) (MA 9681/1-1). Parts of this work were funded by the Deutsche Forschungsgemeinschaft (DFG) – CRC 1450 – 431460824. H.H. thanks the Foundation for Food Safety and Consumer Protection (Stiftung für Lebensmittelsicherheit und Verbraucherschutz) for their generous financial support.

