## Supplemental Information for "Multimodal investigation of endometriosis by using LA-ICP-MS and QCL-based infrared microscopy"

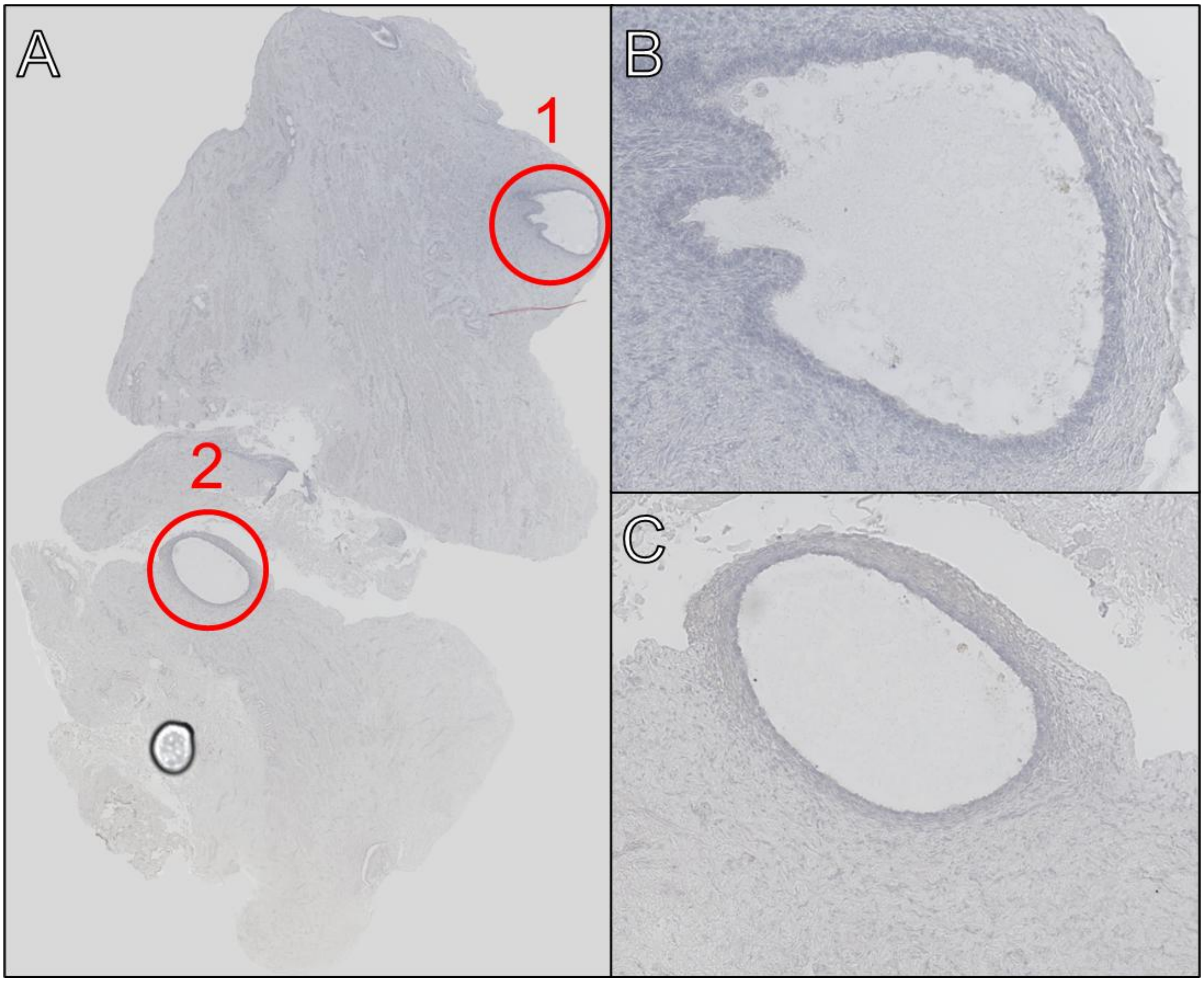


**Figure 1. Hematoxylin and eosin stain of PEM sample**

A) Brightfield microscopic image of a hematoxylin-stained peritoneal endometriosis sample, with red marks to highlight the lesions. B) Zoom in on Lesion 1. C) Zoom in on Lesion 2.

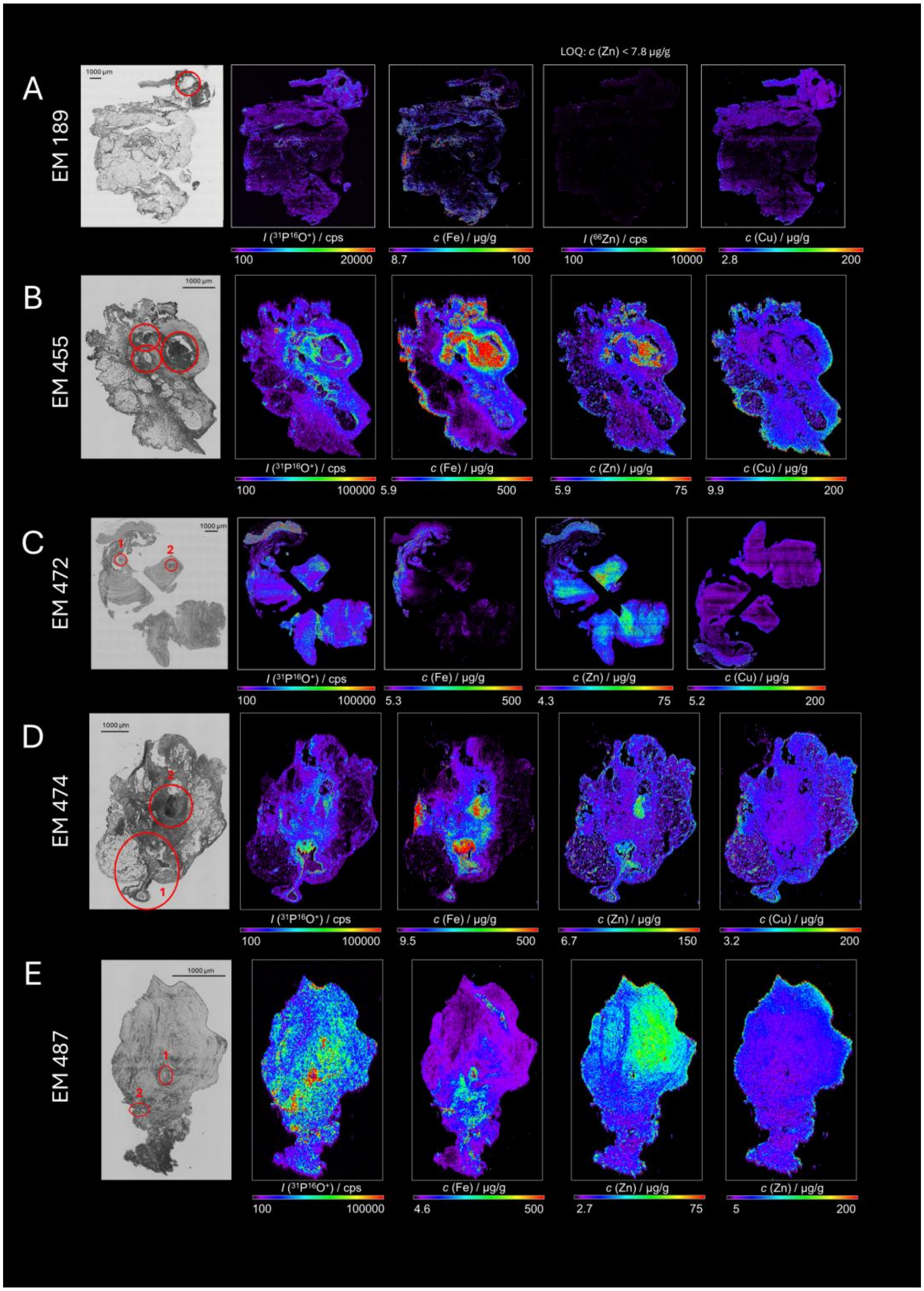


**Figure 2. Elemental distribution of the DIE samples**

A) Brightfield microscopic image of a deep infiltrating endometriosis (sample EM189), with red marks to highlight the lesions. From right to left: elemental Images of P, Fe, Cu, and Zn. The same is shown for sample EM455 (B), sample EM472 (C), sample EM474 (D), and sample EM487 (E) (all deep infiltrated endometriosis).

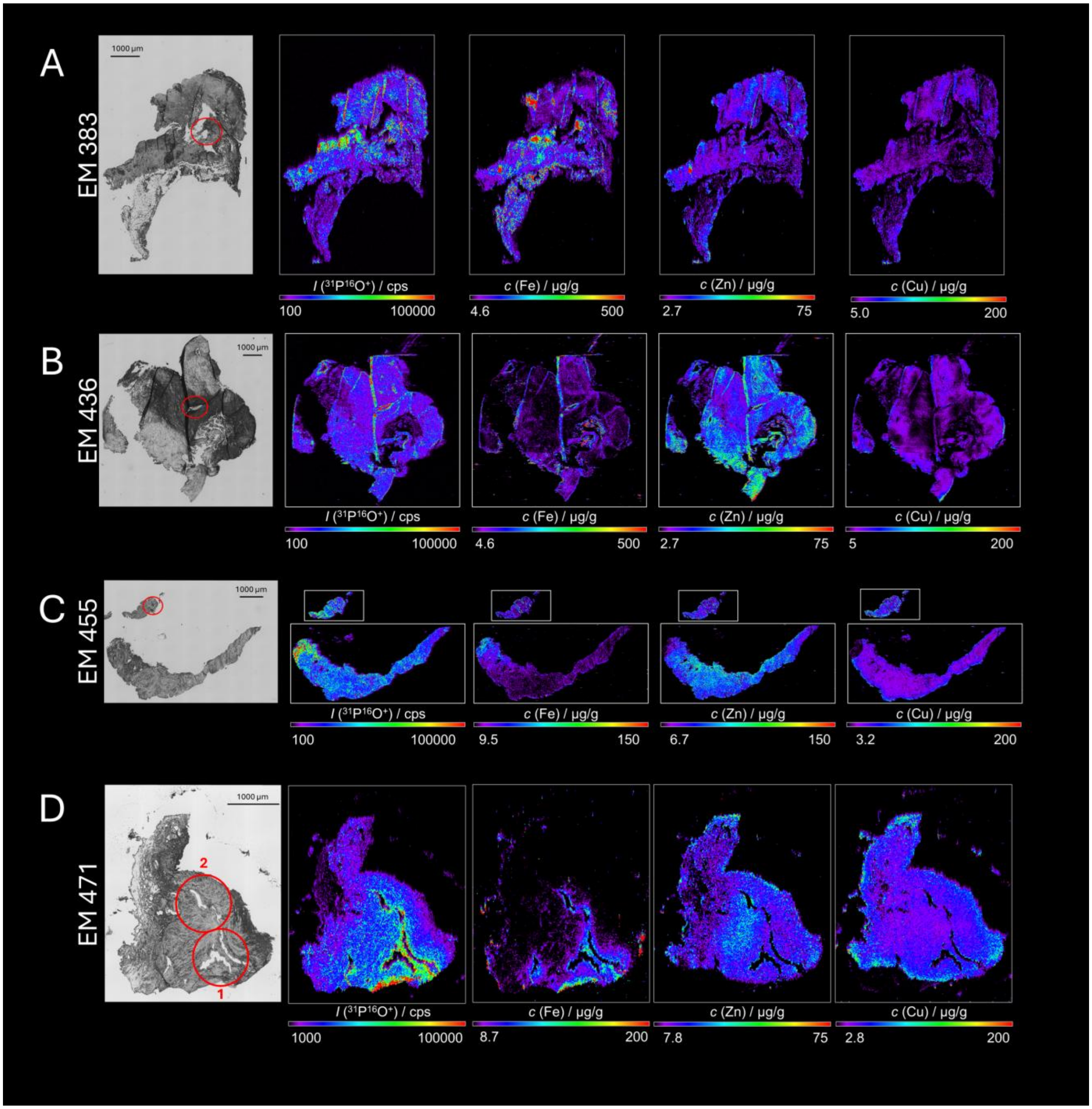


**Figure 3. Elemental distribution of the PEM samples**

A) Brightfield microscopic image of a peritoneal endometriosis (sample EM383), with red marks to highlight the lesions. From right to left: elemental Images of P, Fe, Cu, and Zn. The same is shown for sample EM436 (B), sample EM455 (C), and sample EM471 (D) (all peritoneal endometriosis).

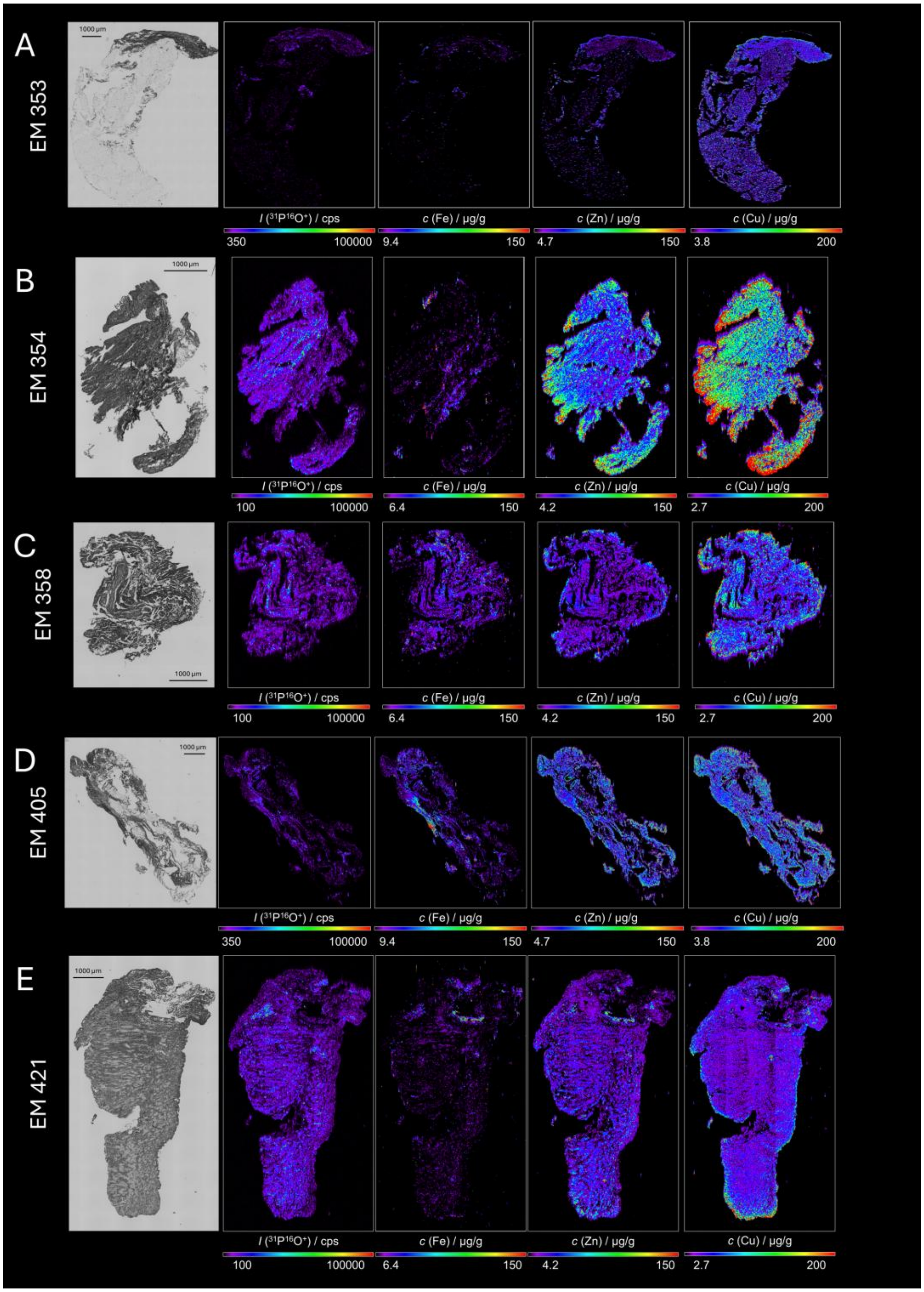


**Figure 4. Elemental distribution of the control samples**

A) Brightfield microscopic image of a control sample (sample EM353), with red marks to highlight the lesions. From right to left: elemental Images of P, Fe, Cu, and Zn. The same is shown for sample EM354 (B), sample EM358(C), sample EM405 (D), and sample EM421 (E) (all control samples).

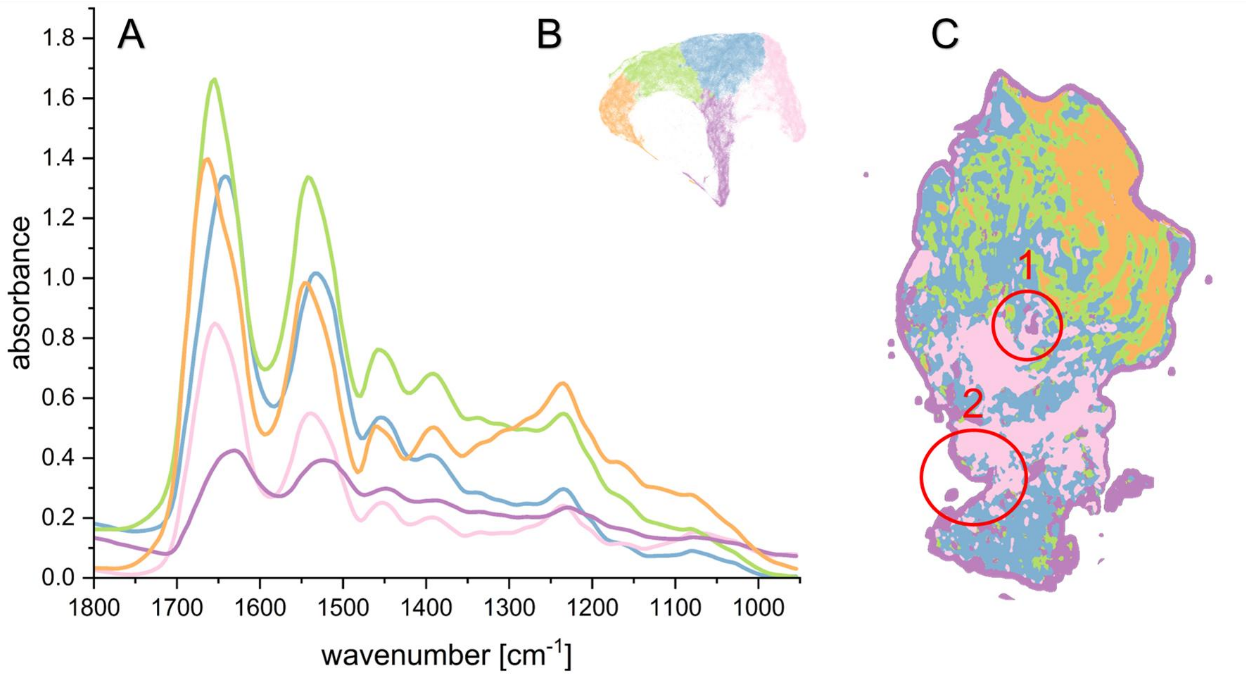


**Figure 5. IR-fingerprint of a DIE lesion.**

A) Average spectra of individual-colored segments of the UMAP embedding of a deep infiltrating endometriosis sample. B) UMAP embedding of the infrared image data. The parameters were set as follows: n_neighbours = 15, min_dist = 0.1 in a cosine metric. The Leiden algorithm was used to color the segments with a resolution parameter of 0.18. IR image, colored according to the segments of B.

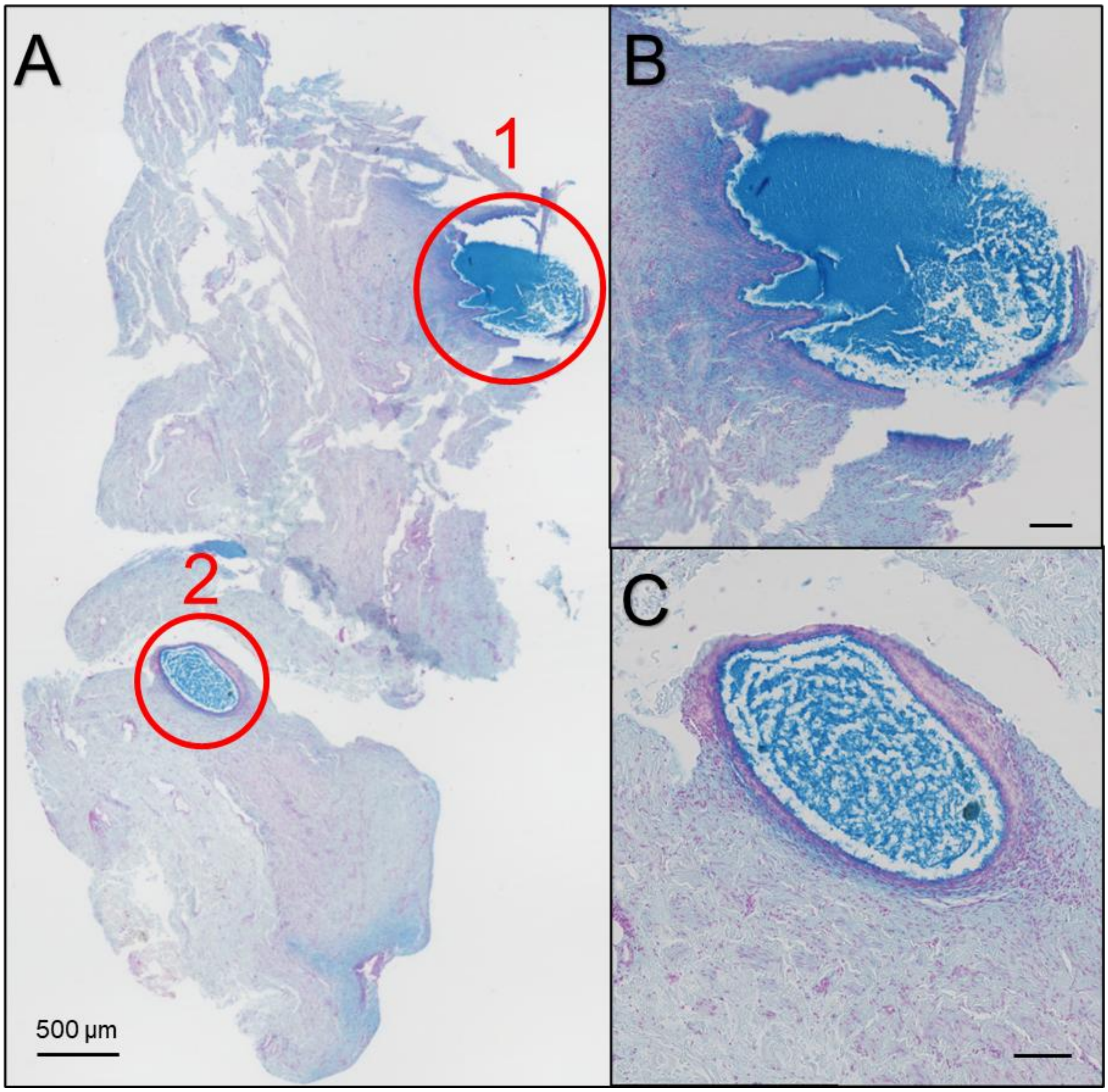


**Figure 6**. **Histological staining of PEM lesion.**

A) Brightfield microscopic image of an alcian blue-stained peritoneal endometriosis sample EM440, with red marks to highlight the lesions. B) Zoom in on Lesion 1. C) Zoom in on Lesion 2.

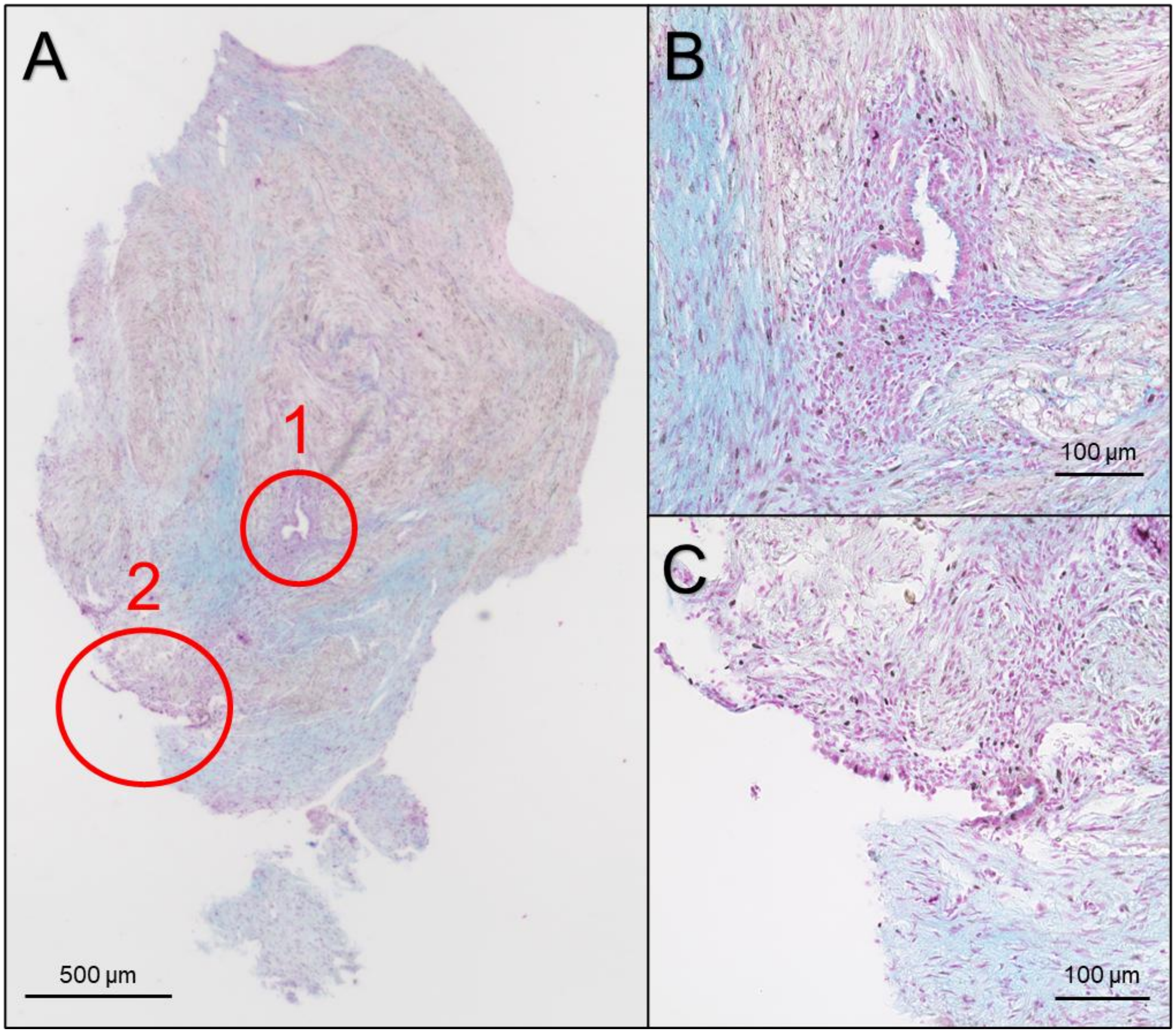


**Figure 7.Histological staining of DIE lesion.**

**A**) Brightfield microscopic image of an alcian blue-stained deep infiltrating endometriosis sample EM487, with red marks to highlight the lesions. B) Zoom in on Lesion 1. C) Zoom in on Lesion 2.
